# Intermittent gross hematuria after lithotripsy: ureteral stent or residual stone?

**DOI:** 10.1101/2020.02.12.20022590

**Authors:** Linjie Peng, Junjun Wen, Wen Zhong

## Abstract

**Purpose:** To explore the role of stone fragment and ureteric stent in process of intermittent gross hematuria (IGH), discuss the definition of ureteral stent related symptoms (USS) in a setting of lithotripsy and clinical outcomes of IGH.

**Methods:** Between January 2018 and July 2019, patients with completed one month follow up after lithotripsy were collected. Based on whether occurrence of IGH, demographics, stent information and clinical outcomes were mainly analyzed.

**Results:** A total of 258 consecutive patients were eventually analyzed, among which 97 patients (37.6%) suffered from IGH. Compared to patients without symptom of IGH, preoperative potassium level (3.89±0.43mmol/L vs. 4.02±0.42mmol/L, p=0.01), initial stone free rate (SFR, 50.52% vs. 68.32%, p=0.007) and potassium citrate application (11.34% vs. 4.55%, p=0.04) were found statistically different in univariate analysis. In multivariate logistic analysis, preoperative potassium level (OR: 0.39, CI: 0.19-0.76, p=0.007), potassium citrate (OR: 2.96, CI: 1.07-8.73, p=0.04), initial SFR (OR: 0.42, CI: 0.24-0.74, p<0.001), and hospital stay (OR: 0.94, CI: 0.89-0.99, p=0.045) were independent risk factors, rather than stent size and stent position. Hemoglobin change, stone area reduction and SFR in one-month follow-up were similar between groups, but more outpatient consultations were found in hematuria group (20 vs. 3, p<0.001), among which 2 patients were readmitted for severe ongoing hematuria.

**Conclusions:** Stone fragment, rather the size or length of ureteral stent, is the independent risk factors of IGH. Definition of USS is not suitable for IGH after lithotripsy, and a comprehensive inform and enough rest would reduce the unnecessary medical consultations.

## Introduction

Hematuria has a prevalence of 4-5% in clinical practice and accounts for 4% to 20% medical consult in urology [1, 2]. The common etiology of hematuria includes urinary stone, coagulation abnormal, trauma, urinary neoplasm and foreign substance in urinary tract. As a worldwide urological issue, approximately 4% to 10% global population suffer from urolithiasis[3-6]. Nearly 90% nephrolithiasis patients went for medical visit with symptom of hematuria[7, 8].

For patients need lithotripsy for active stone removal, endoscopic surgical interventions such as percutaneous nephrolithotomy (PCNL), flexible retrograde intrarenal surgery (RIRS, fURS) and transurethral ureteroscope lithotripsy (URSL) are all well-established [9, 10]. Normally, ureteral stent will be inserted and would stay inside for several weeks to couple months at the end of procedures, which was supposed to provide a better drainage, prevent ureteral obstruction and promote healing of ureteral mucosal at the end of surgery[11, 12]. However, the discomforts followed by ureteral stent are inevitable and disturbing. Intermittent gross hematuria (IGH) during discharge period was commonly classified into ureteral stent related symptoms (USS), together with lumbago, flank pain and other lower urinary tract symptoms (LUTS)[12-15]. Patients who suffer from USS can up to 80%, in which hematuria take up 18.1% to 39.4%[14, 15]. The symptom of IGH would increase the anxiety of patients, which eventually leads to additional outpatient visit after lithotripsy and increases the workload of urologists.

Currently, there are scarce studies focused on USS, neither IGH itself. On the one hand, the role of stone fragment and size and length of ureteral stent had rarely been investigated after lithotripsy. A suitable size or length of ureteral stent was supposed to play a key role in USS, but it were controversial between several studies[16, 17]. To be noticed, only about 60 % to 90% patients attain final stone free, which is differed by ways of lithotripsy. Influence of stone fragment should always be taken into consideration in urolithiasis patients when IGH occurs. But it is still unknown whether IGH is correctly classified into USS, as well as which one plays a major role in setting of lithotripsy. On the other hand, long term IGH would potentially increase outpatient consultation or even readmission. Chronic bleeding and ureteral stent as a foreign substance might increase the risk of urinary tract infection (UTI) or eventually lead to severe anemia.

For a better understanding and provide complementary evidence for further study, this study was designed to study the role of stone fragment and ureteral stent in the occurrence of IGH and the clinical outcomes among patients with indwelling ureteral stent after lithotripsy.

## Materials and Methods

### Study design and participants

This study was approved by the ethics committee of the First Affiliated Hospital of Guangzhou Medical University. Patients met the following criteria between January 2018 and July 2019 in tour center were retrospectively enrolled. Inclusion criteria: 1. Adult patients (18-80 years old) selected endoscopic procedures (PCNL, RIRS or URSL) for urinary stone removal; 2. Discharged with an indwelling ureteral stent after lithotripsy; 3. attained a completely clear appearance of urine for more than one day before discharge; 4. Completed one month follow up after discharge and had complete record of whether occurrence of IGH. Exclusion criteria: 1. Lost follow up, or incomplete data of some medical examination like urine test, urine culture or without CT scan; 2. Severe comorbidities containing severe cardiovascular or respiratory diseases; 3. Receiving anti-coagulation therapy; 4. Patients who declared their data not available for scientific research preoperatively.

The primary study was the incidence of IGH, which was defined as clearly visible red color in urine for more than a week and exclude influence of certain drug or food. Based on the outcome, patients were classified into two groups (Group 0: without IGH, Group 1: with IGH) and related factors supposed to participated in IGH would be further analyzed. The secondary study was following clinical outcomes: change in hemoglobin, stone free rate (SFR) other discomforts or medical consult after discharge. Demographic data, medical information, stent size, stent position, stone size, stone area reduction and SFR in both first postoperative day and first postoperative month were all analyzed.

### Procedures and Follow-up

All the preoperative patient evaluations were performed in accordance with a standard endoscopy suite. None contrast CT (NCCT) scan would be routinely performed prior to the study. *Stone burden* (stone size and area) was assessed as the maximum length or surface area on NCCT. In case of multi-calyx stones, *Stone burden* was the summation of all individual stones. Antibiotic was applied according to the antimicrobial susceptibility test and negative urine culture must be assured before surgery. Procedure flows of PCNL, RIRS and URS were similar to previous studies and guideline recommendation without other interventions[18, 19]. Under general anesthesia, a suitable position was selected for stone removal: prone position for PCNL and supine position for RIRS and URSL. In PCNL, a 4.7-7.0 Fr DJ stent (Cook Urological Inc., Bloomington, IN, USA) was placed antegradely under monitor of x ray at the end of procedure. Retrograde placement and ureteroscope guiding would applied in RIRS and URS. A guidewire was used in all the procedures to assist the insertion of ureteral stent.

To make sure the suitable location of the ureteral stent and assess the status of residual stone, a plain abdominal x ray (KUB) would be routinely performed on first postoperative day, and NCCT would be available in case of radiolucent stone. Stone free was defined as no evidence of residual stones or stone fragments less than 4 mm on radiograph. *A suitable stent position* was supposed that distal curl of ureteric stent did not cross the middle line of human body, or it would be considered too long. Laboratory examinations (blood cell count and chemical items) were also conducted. Some oral medicines supposed to promote a higher stone clearance were advised for patients with residual stone fragments. After a steadily and completely clear appearance of urine was assured, patients were discharged and followed up for symptoms like IGH, lumbago and lower urinary tract symptoms through telephone. Discomforts described above and additional medical consultation or readmission during follow-up would be recorded.

### Statistical analysis

Continuous variables were demonstrated as mean ± SD, and Student’s t-test were used for analysis. Chi-square or Fisher’s exact test was performed in case of categorical variables. Univariate logistic regression analysis was then carried out to identify the significant risk factors using forward selection. Covariates with a p value < 0.2 would be enrolled into multivariate logistic regression analysis subsequently. A two-sided p value < 0.05 was considered statistically significant in the overall outcomes. All statistical analysis was performed using the Statistical Package for the Social Sciences (SPSS), version 13.0 software (SPSS Inc., Chicago, IL, USA).

## Results

As it was shown in Table 1, a total of 258 consecutive patients met the inclusion criteria were retrospectively enrolled into final analysis. The demographics were comparable among two groups. 97 patients (37.6%) suffered from IGH (Group 1) with a mean age of 49.85 years and a mean BMI of 24.19 kg/m^2^. IGH occurred more in males (65.89%), but there no difference was found compared with Group 0 (P=0.15). PCNL, RIRS and URSL were selected for stone removal by 132, 88 and 38 patients, respectively. PCNL seemed to own a higher rate of IGH, compared to RIRS and URS (46.39%, 38.14% and 15.47%, respectively), but no statistical significance was found between two groups (p=0.47). Stone size was 26.17±15.57mm in Group 0 versus 26.32±16.64mm in Group 1 (p=0.94), while stone area was 350.65±477.93mm^2^ and 341.52±569.79mm^2^, respectively (p=0.89). Creatinine level was comparable between two groups (110.76±84.7 vs. 116.24±93.98 mmol/L, p=0.64), but preoperative potassium level was significantly lower among patients with IGH (3.89±0.43mmol/L vs. 4.02±0.42mmol/L, p=0.01).

Ureteric stent would stay for 1 month for all enrolled patients. However, the stent size did not play an important role in gross hematuria compared to patients without IGH (p=0.80), even larger size (≥6Fr) took up a major part (65.98%) in Group 1. Similarly, 44.33% patients in Group 1 showed an inappropriate stent position on radiograph, but no statistical significance was detected when compared with Group 0 (p=0.91). Potassium citrate application after discharge was closely associated with higher incidence of gross hematuria (11.34% vs. 4.55%, p=0.04). To be noticed, highly stone free in 1^st^ postoperative day was attained in Group 0 (68.32% vs. 50.52%, p=0.007), but results of stone compositional analysis were similar between two groups (p=0.95).

As listed in Table 2, stone free rate (73.91% vs. 68.04%, p=0.38) and reduction of stone area were insignificant in 1^st^ postoperative month between groups, even relatively larger area reduction (43.92±51.75mm^2^ vs. 28.25±66.6, p=0.19) was found in Group 1. The change of hemoglobin, platelet and red blood cell and other discomforts like lumbago and LUTS were comparable between two groups. Among patients with gross hematuria, additional outpatient consultation happened in 20 (20.62%) patients, which was much higher (1.86%, p<0.001). only 2 patients (2.06%) in Group 1 were readmitted to hospital because of severe ongoing bleeding.

According to logistic regression analysis (Table 3), size and not suitable position of ureteric stent were not risk factors in IGH (OR 0.9, CI 0.53-1.54, p=0.70 and OR 1.06, CI 0.64-1.77, p=0.81, respectively). Stone free status was significantly different in univariate (OR 0.47, CI 0.28-0.79, p=0.004) and was an independent risk factor for gross hematuria in multivariate logistic analysis (OR 0.42, CI 0.24-0.74, p<0.001). Preoperative potassium level (OR 0.39, CI 0.19-0.76, p=0.007), potassium citrate application after discharge (OR 2.96, CI 1.07-8.73, p=0.04) were both the risk factors in gross hematuria. Specially, hospital stay was showed as a risk factor in multivariate analysis (OR 0.94, CI 0.89-0.99, p=0.045) but not univariate analysis.

## Discussion

Without timely and proper treatments for urolithiasis, hydronephrosis, urinary infection and chronic kidney disease are chronic threat to health and even mortality, which are great economic burden to patients[20]. To treat urolithiasis, ureteric stent is routinely inserted after lithotripsy. However, the longer ureteral stent staying means the higher possibility of discomforts. Higher rate of discomforts occurred means additional medical counseling or even readmission needed, which is a great disturbance for patients and increases the workload of urologists. But severe complications like complexed urinary infection, severe blood loss needing blood transfusion followed by intermittent gross hematuria were not recorded.

It used to believe that friction between urinary tract and ureteric stent accounts for IGH. At the same time, IGH was ambiguously classified into USS without specific definitions. Several studies about USS and ureteric stent were controversial in terms of the size and length of ureteric stent[16, 21-23]. However, even patients were discharged in our hospital after adequately, who were suggested to take adequate rest before stent were removed, 37.6% of them still suffered from IGH. In present study, we did not have enough evidences to support the size and the position of stent participated in IGH. When the confounders like daily activities and ureteral stent were excluded, there must be something else involving in this process. As found in present study, SFR was significantly lower in IGH group in first postoperative day. However, the discrepancy turned insignificant in first postoperative month, which meant the passage of stone fragment played an indispensable role in IGH. In addition, the stone area reduction in patients without stone free between postoperative 1^st^ day and 1^st^ month had proved the stone passage after discharge. Therefore, stone free statue in postoperative 1^st^ day was an independent risk factors and patients achieved stone free were less likely to suffer from IGH after lithotripsy. On the other hand, the definition of USS need to be more specific and it is inappropriate using USS to explain IGH in case of lithotripsy.

The activities of discharge patients cannot be controlled, but a longer hospital stay may be possible, as we found a longer hospital stay was a protective factor in IGH. Owing to a less intensity of daily activities, a longer hospital staying meant a longer time for urinary tract recovery. To some extent, it also explained the reason why male patients took up a larger portion in IGH, rather than females in another aspect. Even a clear urine without obvious gross hematuria should be assured before patients discharged, but certain patients with higher risk of IGH should be informed of the possibility of IGH and necessity of enough rest.

Hypocitraturia is widely known as a risk factor for the development and recurrence of urolithiasis and is present in 20–60% of patients[24]. Potassium citrate alkalinizes the urine and solubilizes urine calcium, reducing the risk of urolithiasis recurrence, particularly in those found to have low urine citrate [25, 26]. In present study, potassium citrate was applied in 7 patients without stone free but none of them attain stone free in 1^st^ month, which was consistent with previous study[27]. However, 11 patients with stone free and potassium citrate application suffered from IGH, which might suggest not only passage of stone fragment involved but also the metabolic change mattered. And stent might have subtle influence too, even no significant difference was found.

Potassium as another item of metabolic, is one of main cations that maintains body’s normal osmotic pressure and acid-base balance, participating in sugar and protein metabolism, and ensuring the normal function of nerve muscles. Both hyperkalemia and hypokalemia are thought to be associated with increased morbidity and mortality among hospitalized patients. In our study, we found a relative low level of serum potassium was associated with higher occurrence of gross hematuria, even they were in the normal range of internal environment. For a better understanding, we analyzed the difference between relatively rower and higher concentration of potassium level (3.98 mmol/L). We found male patients were significantly more in the IGH group (94 VS. 76, p=0.009), which might explain this phenomenon to some extent as we discussed before. Additionally, the application of potassium citrate was to supplement the level of potassium in hypokalemia patients. In all, a better understanding of potassium level and potassium citrate in the process of IGH require further basic researches.

Some limitations should be noticed when this study’s findings are interpreted by clinicians. First of all, a retrospective review is an inherent limitation. Secondly, as a tertiary medical center with a lot of complicated urolithiasis, our patient population might not represent general urology patients. Further prospective randomized controlled trials are welcomed to testified what were found in present study.

## Conclusions

Occurrence of IGH after lithotripsy being classified into ureteral stent related symptoms is inappropriate, because stone fragment plays a major role in IGH but not ureteric stent. Together with SFR in 1^st^ postoperative day, preoperative level of serum potassium, application of potassium citrate and hospital stay, SFR in postoperative 1^st^ morning was independent risk factors in the process of IGH. There was no severe blood loss, but more outpatient consultation was found in IGH group.

## Data Availability

Availability of all data referred to in the manuscript were included as supplementary documents.

## Author Contributions

ZW had full access to all the data in the study and takes responsibility for the integrity of the data and the accuracy of the data analysis. PLJ owned the study concept and design. WJJ contributed to data collection. ZW and PLJ performed statistical analysis and drafted the manuscript. ZW was responsible for critical revision of the manuscript for important intellectual content.

## Informed consent

Informed consent was obtained from all individual participants included in the study.

## Abbreviations

IGH: intermittent gross hematuria
USS: ureteral stent related symptoms
LUTS: lower urinary tract symptoms
PCNL: percutaneous nephrolithotomy
RIRS: flexible retrograde intrarenal surgery
URSL: transurethral ureteroscope lithotripsy

## Reference

1. Bolenz C, Schroppel B, Eisenhardt A, Schmitz-Drager BJ, Grimm MO: The Investigation of Hematuria. Deutsches Arzteblatt international 2018, 115(48):801–807.

2. Avellino GJ, Bose S, Wang DS: Diagnosis and Management of Hematuria. The Surgical clinics of North America 2016, 96(3):503–515.

3. Pfau A, Knauf F: Update on Nephrolithiasis: Core Curriculum 2016. Am J Kidney Dis 2016, 68(6):973–985.

4. Stamatelou KK, Francis ME, Jones CA, Nyberg LM, Curhan GC: Time trends in reported prevalence of kidney stones in the United States: 1976-1994. Kidney Int 2003, 63(5):1817–1823.

5. Zeng G, Mai Z, Xia S, Wang Z, Zhang K, Wang L, Long Y, Ma J, Li Y, Wan SP et al: Prevalence of kidney stones in China: an ultrasonography based cross-sectional study. BJU international 2017, 120(1):109–116.

6. Hesse A, Brandle E, Wilbert D, Kohrmann KU, Alken P: Study on the prevalence and incidence of urolithiasis in Germany comparing the years 1979 vs. 2000. Eur Urol 2003, 44(6):709–713.

7. Mayans L: Nephrolithiasis. Primary care 2019, 46(2):203–212.

8. Gottlieb M, Long B, Koyfman A: The evaluation and management of urolithiasis in the ED: A review of the literature. Am J Emerg Med 2018, 36(4):699–706.

9. Turk C, Petrik A, Sarica K, Seitz C, Skolarikos A, Straub M, Knoll T: EAU Guidelines on Interventional Treatment for Urolithiasis. European urology 2016, 69(3):475–482.

10. Assimos D, Krambeck A, Miller NL, Monga M, Murad MH, Nelson CP, Pace KT, Pais VM, Jr., Pearle MS, Preminger GM et al: Surgical Management of Stones: American Urological Association/Endourological Society Guideline, PART II. The Journal of urology 2016, 196(4):1161–1169.

11. Makarov DV, Trock BJ, Allaf ME, Matlaga BR: The effect of ureteral stent placement on post-ureteroscopy complications: a meta-analysis. Urology 2008, 71(5):796–800.

12. Shen PF, Li YT, Yang J, Wei WR, Dai Y, Zeng H, Wang J: The Results of Ureteral Stenting After Ureteroscopic Lithotripsy for Ureteral Calculi: A Systematic Review and Meta-Analysis. J Urology 2011, 186(5):1904–1909.

13. Lamb AD, Vowler SL, Johnston R, Dunn N, Wiseman OJ: Meta-analysis showing the beneficial effect of alpha-blockers on ureteric stent discomfort. Bju Int 2011, 108(11):1894–1902.

14. Joshi HB, Okeke A, Newns N, Keeley FX, Jr., Timoney AG: Characterization of urinary symptoms in patients with ureteral stents. Urology 2002, 59(4):511–516.

15. Wang Y, Xu M, Li W, Mao Y, Da J, Wang Z: It is efficient to monitor the status of implanted ureteral stent using a mobile social networking service application. Urolithiasis 2019.

16. Calvert RC, Wong KY, Chitale SV, Irving SO, Nagarajan M, Biyani CS, Browning AJ, Young JG, Timoney AG, Keeley FX et al: Multi-length or 24 cm ureteric stent? A multicentre randomised comparison of stent-related symptoms using a validated questionnaire. BJU international 2013, 111(7):1099–1104.

17. Damiano R, Autorino R, De Sio M, Cantiello F, Quarto G, Perdona S, Sacco R, D’Armiento M: Does the size of ureteral stent impact urinary symptoms and quality of life? A prospective randomized study. Eur Urol 2005, 48(4):673–678.

18. Patel U, Anson KM: Percutaneous Nephrolithotomy Made Easier: A Practical Guide, Tips and Tricks. Bju Int 2008, 102(9):1178–1178.

19. Breda A, Ogunyemi O, Leppert JT, Schulam PG: Flexible ureteroscopy and laser lithotripsy for multiple unilateral intrarenal stones. Eur Urol 2009, 55(5):1190–1196.

20. Sur RL: Primary Prevention of Nephrolithiasis Is Cost-Effective for a National Healthcare System Editorial Comment. Bju Int 2012, 110(11c):E1068–E1068.

21. Cubuk A, Yanaral F, Ozgor F, Savun M, Ozdemir H, Erbin A, Yuksel B, Sarilar O: Comparison of 4.8 Fr and 6 Fr ureteral stents on stent related symptoms following ureterorenoscopy: A prospective randomized controlled trial. The Kaohsiung journal of medical sciences 2018, 34(12):695–699.

22. Nestler S, Witte B, Schilchegger L, Jones J: Size does matter: ureteral stents with a smaller diameter show advantages regarding urinary symptoms, pain levels and general health. World J Urol 2019.

23. Fischer KM, Louie M, Mucksavage P: Ureteral Stent Discomfort and Its Management. Current urology reports 2018, 19(8):64.

24. Domrongkitchaiporn S, Stitchantrakul W, Kochakarn W: Causes of hypocitraturia in recurrent calcium stone formers: Focusing on urinary potassium excretion. American Journal of Kidney Diseases 2006, 48(4):546–554.

25. Carvalho M, Erbano BO, Kuwaki EY, Pontes HP, Liu JWTW, Boros LH, Asinelli MO, Baena CP: Effect of potassium citrate supplement on stone recurrence before or after lithotripsy: systematic review and meta-analysis. Urolithiasis 2017, 45(5):449–455.

26. Soygur T, Akbay A, Kupeli S: Effect of potassium citrate therapy on stone recurrence and residual fragments after shockwave lithotripsy in lower caliceal calcium oxalate urolithiasis: a randomized controlled trial. J Endourol 2002, 16(3):149–152.

27. Kang DE, Maloney MM, Haleblian GE, Springhart WP, Honeycutt EF, Eisenstein EL, Marguet CG, Preminger GM: Effect of medical management on recurrent stone formation following percutaneous nephrolithotomy. J Urology 2007, 177(5):1785–1788.

